# “Emotional stress is more detrimental than the virus itself”: Towards an understanding of HIV testing and pre-exposure prophylaxis (PrEP) use among internal migrant men in South Africa

**DOI:** 10.1101/2023.08.23.23294421

**Authors:** Maria F. Nardell, Caroline Govathson, Salomé Garnier, Ashley Watts, Dolapo Babalola, Nkosinathi Ngcobo, Lawrence Long, Mark N. Lurie, Jacqui Miot, Sophie Pascoe, Ingrid T. Katz

**Author notes:** Corresponding author: Maria Francesca Nardell. Equal contribution (Sophie Pascoe, Ingrid T. Katz).

## Abstract

**Introduction:** South Africa has one of the highest rates of internal migration on the continent, largely comprised of men seeking labor in urban centers. South African men who move within the country (internal migrants) are at higher risk than non-migrant men of acquiring HIV yet are less likely to test or use pre-exposure prophylaxis (PrEP). However, little is known about the mechanisms that link internal migration and challenges engaging in HIV services.

**Methods:** We recruited 30 internal migrant men (born outside Gauteng Province) for in-depth qualitative interviews at sites in Johannesburg (Gauteng) where migrants may gather (i.e., factories, homeless shelters). Interviewers used open-ended questions, based in the Theory of Triadic Influence, to explore experiences and challenges with HIV testing and/or PrEP. A mixed deductive inductive content analytic approach was used to review data and explain why participants may or may not use these services.

**Results:** Migrant men come to Johannesburg to find work, but their struggle to survive without reliable income causes daily stress. Stress and time constraints limit their availability to seek health services, and many men lack knowledge about the opportunity for PrEP should they test negative. In addition, migrants must also adjust to life in Johannesburg, where they may be unfamiliar with where to access HIV services and lack social support to help them do so. Migrants may also continue to travel intermittently for work or social purposes, which can make it hard to take a daily pill like PrEP. Yet Johannesburg also presents opportunities for HIV services for migrant men, especially those who perceive greater availability and anonymity of HIV information and services in the city as compared to their rural homes of origin.

**Conclusions:** Bringing HIV services to migrant men at community sites may ease the burden of accessing these services. Including PrEP counseling and services alongside HIV testing may further encourage men to test, particularly if integrated into counseling for livelihood and coping strategies, as well as support for navigating health services in Johannesburg.

## Introduction

Despite substantial progress, South Africa continues to have one of the world’s largest HIV epidemics, with approximately 7.5 million people living with HIV.^1^ While women shoulder a greater burden of HIV than men in South Africa,^2^ there are major gaps in reaching men with HIV services.^3–6^ In particular, it is estimated that only half of men living with HIV in South Africa know their status,^3,7^ and levels of HIV testing for men remain consistently lower than those for women.^8–10^ Men with HIV also have lower rates of treatment adherence, viral suppression, and survival.^3,11,12^ Alongside efforts to help men living with HIV to start and stay on treatment, it is imperative to focus on testing and prevention for men not already infected.^13^ Oral pre-exposure prophylaxis (PrEP) is highly effective at preventing HIV.^14–17^ South African men have shown interest in using PrEP,^18,19^ but barriers to men’s use of HIV services include masculine norms which discourage care seeking, anticipated stigma in health facilities, poverty and unemployment, and lack of knowledge about PrEP.^3,20^ In addition, most PrEP services in South Africa have targeted women, men who have sex with men, and sex workers.^21–24^ Concerns about the cost-effectiveness of providing PrEP to heterosexual men may limit its implementation.^25^ Despite this, South Africa’s National Strategic Plan for HIV, TB, and STIs 2017–2022 recommends PrEP for anyone who reports being at risk for HIV, including heterosexual men.^26^ Thus, it is important to understand how to target PrEP delivery to men for whom it may provide greatest benefit.

South African men who move within the country, called internal migrants,^27,28^ bear as much as two to three times the burden of HIV as compared to non-migrant men in South Africa,^29,30^ as well as a higher burden than international migrants.^30^ Associations between HIV vulnerability and migration are complex and vary among migrant groups.^31^ Some studies have shown that migrant men are at higher risk for HIV acquisition during the disruptive process of relocation^32^ and engage in higher sexual risk behaviors.^33^ Other research has shown that men living with HIV are more likely to become migrants than those who are HIV negative, in part due to marital dissolution, which people living with HIV are more likely to experience.^27^ Compared to non-migrants, internal migrants are less likely to use health services overall, more likely to use private services or traditional healers, and less likely to test for HIV.^34,35^ Migrants are identified as a “key population” for HIV prevention,^31^ but only 15% internal migrant men surveyed in community sites in Johannesburg were aware of PrEP.^30^ HIV interventions that seek to engage men in testing and prevention, such as workplace programs and partner notification strategies,^36^ may exclude men who are unemployed or unpartnered, as is common for migrant men.^30^ Moreover, large trials to improve HIV testing uptake among men in South Africa have found substantial challenges reaching men in communities with high mobility and in-migration.^37^ South Africa has one of the highest rates of internal migration on the continent^38^ with an estimated net migration of a million internal migrants settling in urban Gauteng Province from 2016 to 2021 alone, dwarfing the number of international immigrants by as many as five to one.^39^

Research to unpack the relationship between migration, masculinity, and barriers to HIV testing and prevention in South Africa has been limited. In particular, there is a lack of research on internal migrants’ experience with HIV care, for whom factors such as xenophobia within the healthcare system and lack of knowledge of the South African health care system may not apply.^40–42^ In this paper, we seek to fill this gap by exploring the psychosocial and structural factors that influence HIV testing and prevention uptake among internal migrants through qualitative interviews with internal migrant men in Johannesburg, South Africa. South Africa must engage this key population if it is to meet its national goal of eliminating HIV as a public health threat with “nobody left behind.”^43^

## Methods

### Study Setting and Participants

This study is based in Johannesburg, the most populous city in South Africa. In our team’s prior pilot work among migrant men in Johannesburg, we chose community-based recruitment sites in the Hillbrow, Midrand, Woodmead and Roodeport areas following a period of observation and discussion with community stakeholders. For this study focusing specifically on internal migrant men, we selected two sites where there were high proportions of internal migrant men, a site in Midrand area north of Johannesburg close to factories and the Displaced Persons Unit (DPU) in Hillbrow (Johannesburg central.) Similar to our prior research among this population,^30,44,45^ we chose to focus on community sites in order to reach men who may not seek care at clinics.

Recruitment occurred in August 2022. Team members recruited participants by first asking leadership at the sites if the study team could recruit there on specific days. On site, the research assistants (RA) distributed study flyers. These flyers gave information about the study and invited men to contact the study team if they were interested in participating. Those men who made contact were then given further information and taken through the consent process in-person. The team checked telephone numbers, names, and identifiers to ensure that the same participant did not enroll twice.

### Eligibility and Sampling

We included men who were 18 years or older, self-reported as an internal migrant (defined as a South African born outside of Gauteng Province), were willing to participate in a 45 to 60-minute interview, and able to provide informed consent. Men were excluded from the study if they were believed to be intoxicated at the time of consent (in which case, they were invited to return the following day), unable to understand the study information, or had previously enrolled in the study. In addition to these eligibility criteria, we purposively sampled to seek perspectives both from migrants who reported living in Johannesburg for a year or less, and those who lived in Johannesburg for longer durations.

### Study Design Conceptual Framework

Interviews followed an in-depth semi-structured interview guide developed to explore internal migrant men’s experiences, knowledge, and attitudes towards HIV testing and prevention. The development of our guide was informed by the Theory of Triadic Influence (TTI), a theoretical model of health behavior used in several HIV-related studies, including in South Africa and by our team previously.^46–48^ TTI posits that there are three streams of influence on health-seeking behavior: *individual factors* (e.g., personal motivations, knowledge), *social factors* (e.g., interpersonal relationships, social norms), and *structural factors* (e.g., healthcare system structures and policies). Interview questions examined attitudes and behaviors toward HIV testing and PrEP within each of these streams of influence, such as knowledge of PrEP (*individual factors*), anticipated HIV stigma (*social factors*), and access to healthcare services (*structural factors*).

### Data Collection and Preparation

Qualitative interviews were completed in person by one of three RAs in a private, secure setting in August 2022. (See Interview guides in Appendix.) RAs were trained in qualitative interviewing and fluent in isiZulu, isiXhosa, English and seSotho so as to be able to conduct the interview in the participant’s preferred language. Interviews lasted 45 to 60 minutes. After completing the survey, participants were reimbursed for their time with a ZAR150 (∼USD10) electronic shopping voucher, sent directly to the participant’s cell phone number. The RA conducting the interview recorded it and took notes. All interview recordings were sent to a third-party transcription service who transcribed it, and if in a language other than English, translated it. Each interview was reviewed by the RA who conducted the interview to ensure accurate transcription and translation. Transcripts were assigned a unique study identification number to protect participant confidentiality.

### Data Analysis

Analysis of data aimed to characterize participants’ reasons for engaging or not engaging with HIV testing and PrEP services. We used a mixed deductive and inductive approach informed by grounded theory.^49^ Two team members (MFN, SG) developed an initial codebook based on key concepts from TTI and the interview protocol, as well as emergent codes from a review of the initial five interviews. The coding team (MFN, SG, DB, AW) then discussed emerging themes through repeated review of the transcripts and memoing. Through weekly meetings, the coding team further refined the codebook, resolving discrepancies through discussion, maintaining an audit trail of notes and each codebook version. Interviews were coded in Dedoose.^50^ Each coding team member coded the first 20% of transcripts, and interrater reliability testing resulted in a pooled Cohen’s kappa of 0.80. The four coders individually coded the remaining transcripts. We then reviewed excerpts for each code, identifying repeated patterns of content which formed the basis for broader themes.^49^

### Ethical Approval

The ethics committees at the University of the Witwatersrand (M191068), Mass General Brigham (Harvard University) (2020P002251), and Boston University (H-40529) approved the study. All participants provided written informed consent.

## Results

### Participant Characteristics

We randomly recruited 30 participants for this study, half at the Midrand factories site and half at the Hillbrow homeless shelter. The demographic information for these participants is shown in Table 1. Nearly a third (30%) of participants were born in Limpopo Province, followed by nearly a fifth (16.7%) in KwaZulu-Natal. Almost half (46.7%) of participants spoke isiZulu as their primary language and came to Johannesburg looking for work. Most reported no regular intimate or married partner (66.7%), though half (50%) had children. Nearly half (43.3%) of participants reported living in Johannesburg for less than three years, and nearly a fourth (23.3%) for less than six months. Half (50.0%) of participants reported traveling to see family. Most participants had no regular form of employment. One participant reported living with HIV.

**Table 1.**
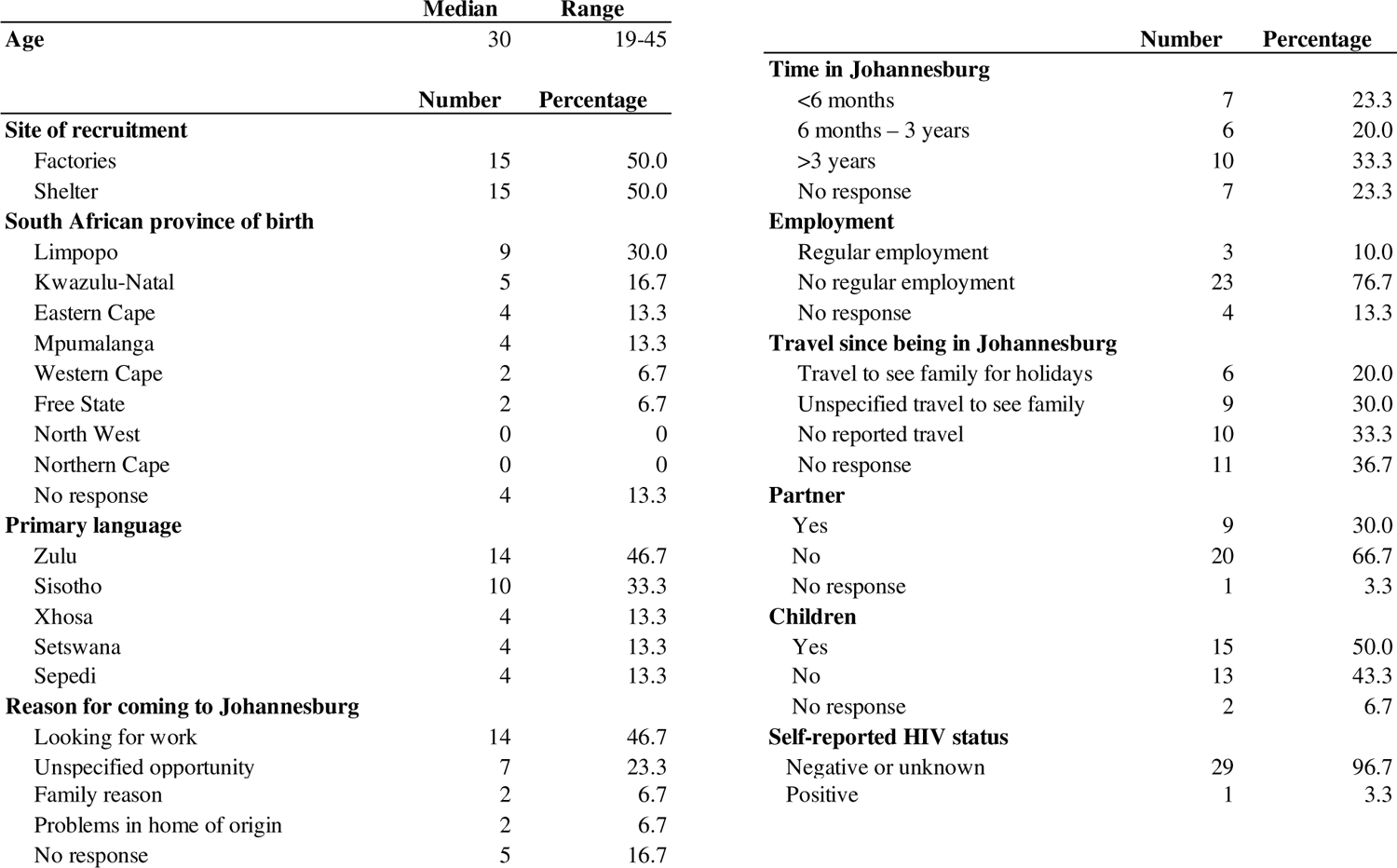
Participant Characteristics. Socio-demographic characteristics of 30 participants.

### Qualitative Results

We organized themes into barriers to and opportunities for care engagement. Within each of these sections, we identified and organized factors at individual, social, and structural levels. Within each of these levels, we identified factors that apply to men more generally as well as factors specific to migrants.

### Individual Barriers

Participants framed their decisions around HIV testing within the context of the high levels of stress they already experience on a day-to-day basis. As shown in Table 2, migrant men described a “weight” over their lives “that is really heavy from problems and life situations,” including “depressing thoughts” about finding work. Adding to their stress was their awareness of HIV risk from unprotected sexual encounters. For most participants, this risk awareness was not a motivator to seek HIV testing. Rather, HIV testing was more often perceived as an added burden because of fear that a positive result “might be too stressful for them.” While almost all participants were knowledgeable about the fact that ART can manage HIV, they nonetheless described the possibility of a positive diagnosis in dire terms, including “about to die,” “ruin my life,” “like suicide,” “painful,” and “make [me] more sick.” As one participant described, “We as men are already carrying a lot. To add another weight on top of that… Emotional stress is more detrimental than the virus itself.” This burden was further exacerbated for many participants by virtue of their move to Johannesburg. Migrant men described challenges with navigating a new city, finding health services, and sometimes personal and family challenges that preceded their move to Johannesburg, which further taxed their cognitive and emotional bandwidth.

**Table 2.** Individual-level barriers to HIV testing and PrEP use.

| Barriers | Examples |
| --- | --- |
| Challenges coping with life stressors | "Naturally men have that weight over them that is really heavy from problems and life situations. We know that it is in our nature to always make mistakes. Knowing that I have to test is going to put on more weight on me. I feel it will be adding to the problems to go and test. It is going to give me depression to find out that I am HIV positive. It is not that we do not want to test. It's just that we do not want to know at that certain time." (Migrant from Western Cape) |
| Fears of testing HIV positive | "People are scared to test. They will rather live in fear not knowing then to find out they are about to die." (Migrant from Limpopo) |
| Lack awareness or knowledge of PrEP | "I do not know about [PrEP]. I have only heard rumours. I have never come across someone who have taken them." (Migrant from Mpumalanga) |
| Seek healthcare for symptoms | "When I need health care I go to a public clinic because it is free. I wait and see if the sickness lasts for 48 hours then I go to the clinic. Anything that lasts for less than that I can handle." (Migrant from Mpumalanga) |

Complicating this narrative was limited awareness about PrEP among most (60%) participants. Even those who had heard of it had little knowledge of anyone who had used it before, where to find it, or how to use it. Participants also mentioned that awareness of HIV prevention options was particularly low in rural areas of the country, where many migrants had been raised. Given most participants did not know PrEP would be an option if they tested negative, HIV testing was associated with few upsides. Thus, fears of testing positive outweighed the potential for relief if they tested negative. While all participants reported testing for HIV at least once in their life, most tested only when required by a healthcare provider, rather than for prevention. The interviewer’s mention of PrEP was usually met with interest, with several respondents mentioning how they would trust PrEP if it came from healthcare professionals and after they “see it works for certain individuals.”

### Social Barriers

Gender norms played a large role in influencing participants’ beliefs and behaviors around HIV testing and PrEP. Participants described how HIV testing was felt to be a woman’s responsibility, and a female partner’s status was assumed to reflect their own status as well. As shown in Table 3, most participants reported that clinics are designed to serve women’s needs, and the predominance of female health care providers in clinics could be a disincentive to seeking care. Some migrants described how traditional gender norms were particularly common in their rural homes of origin, and it was hard to shift this perspective in Johannesburg. In addition, social gatherings with other men, particularly involving alcohol, emerged as a barrier to PrEP use. Participants described how men “are most vulnerable” when they drink because it distracts them from taking pills. Drinking also increases their HIV risk because men are less likely to use condoms when intoxicated.

**Table 3.** Social-level barriers to HIV testing and PrEP use.

| Barriers | Examples |
| --- | --- |
| Social gender norms | "Women do not have such problems because the clinics are created for them. Women bear children. So already, they are used to the clinic. Women have many illnesses. I think it is women that transfer infections to men. Maybe going to a men's clinic would be better. It is quite a challenge to be seen by women going to the clinic." (Migrant from Western Cape) |
| Social alcohol use | "It is really difficult to help men. Most men like to drink....Being drunk can make them miss the pill if it to be taken daily." (Migrant from KwaZulu-Natal) |
| Anticipated stigma |  |
| Testing HIV positive | "You worry that when you go for an HIV test your results might be different and that will affect the relationship negatively. I think that knowing your status puts a lot of pressure on you." (Migrant from Eastern Cape) |
| PrEP | "I think the problem might be to take [PrEP] pills in front of other people." (Migrant from Eastern Cape) |
| Travel and migration | "Different people, different places. You must be prepared to be misunderstood [when traveling]." [Migrant from Free State] |

Anticipated stigma was a major concern reported by nearly all men, including stigma from clinic staff and within communities. Although most participants reported that they themselves were comfortable with others having HIV, they nonetheless worried that testing positive would negatively affect their relationship with partners, family, or friends, who “discourage each other from testing.” PrEP use posed additional concerns, including the fear that PrEP would be mistaken for ART or signal infidelity. In addition, a few participants who knew of PrEP mentioned that PrEP is associated with men who have sex with men (MSM), which prompted some men “to shy away from using it.” Although this study focused on migrants within South Africa, one participant described himself as “a foreigner” within Johannesburg, even though he had been in the city for 12 years, which raised concerns for him about how he would be treated in healthcare spaces. Another participant felt that travel and being a newcomer in Johannesburg for less than a year increased the risk for being “misunderstood” by people who did not know him.

### Structural Barriers

Structural factors emerged as the largest barrier to HIV testing and PrEP use for migrant men. Finding and maintaining work was the biggest priority for all participants. Some managed to find intermittent “piece jobs,” i.e., unpredictable day jobs in which men are paid by the hour or day for the work that is done. Others described how their jobs were unexpectedly terminated or how their pay was insufficient to afford housing or food. Given pressures to work, time was a major barrier to HIV testing, especially at clinics where participants reported queuing all day. As shown in Table 4, one participant said, “I have to spend the entire day on the street…waiting for an opportunity for piece work. If I go for testing, I might miss that opportunity.” Many migrants reported coming to Johannesburg given its reputation as the “City of Gold” where “opportunities are broad.” For them, the gap between their dreams and reality was especially stark. Participants reported that work was not just a means of survival but the necessary precursor to “a home, wife, and kids…a dream [which] has not materialized.” On the other hand, for those participants with a family, the immediate need to provide for their children emerged as a higher priority than the time or transportation costs involved for HIV testing.

**Table 4.** Structural-level barriers to HIV testing and PrEP use.

| Barriers | Examples |
| --- | --- |
| Employment opportunity costs | "I have to spend the entire day on the street by the robot waiting for an opportunity for piece-work. If I go for testing I might miss that opportunity." (Migrant from KwaZulu-Natal) |
| Traveling for work | "Because I am always on the go. Even with the piece-jobs sometimes I don't sleep at home. I am concerned that I might miss a pill." (Migrant from Limpopo) |
| Adjustment to Johannesburg | "I am still new in Johannesburg. It is a busy city. It is full of criminals. You cannot even ask people for directions to the clinic. Sometimes health facilities are located down town where we cannot go because it is dangerous. Those are the challenges." (Migrant from Mpumalanga) |
| Lack of residency documentation | "The issue of illegal immigrants is a serious problem in South Africa. We have many illegal immigrants in the country. You find people sick they cannot access health services because they are here illegally. I feel that clinics should be open even to illegal immigrants. If a person has a passport, they should be able to access health services." (Migrant from Limpopo) |

On top of their livelihood concerns, migrants described challenges adjusting to life in Johannesburg, including concerns about the fast pace of the city and their own safety. Participants also described how migrants may be unfamiliar with where HIV services are located, lack community support to access health services, or not trust services from people they don’t know. Migrants reported that travel to other parts of the city or country could increase HIV risk as they were more likely to have unprotected sexual encounters during these times. None of the participants used PrEP currently, though some hypothesized that traveling, including overnight travel for piece jobs, might pose a challenge for taking a daily pill. Another participant also mentioned how “illegal immigrants” have difficulty accessing healthcare in South Africa.

### Opportunities

As shown in Table 5, participants described several opportunities for engagement in HIV testing, prevention, and care, including those that relate to many of the themes above. Despite the challenges of daily life in Johannesburg, participants noted that HIV services in community-based settings such as pharmacies, pop-up tents, and shops made access easier. Most appreciated the sense of anonymity they felt seeking healthcare in Johannesburg as compared to rural areas, where they had more concern for unintentional disclosure from clinic staff or other patients who knew them. In fact, some participants reported that if diagnosed with HIV, they would feel most comfortable seeking support from healthcare staff in Johannesburg. More readily available HIV information in Johannesburg was also perceived as an opportunity to be harnessed to promote greater care engagement. As one participant shared, “I think people need to be informed about PrEP. This will encourage more people to do HIV testing so they can use PrEP if their results are negative.”

**Table 5.** Opportunities for HIV testing and PrEP use.

| **Opportunities** | **Examples** |
| --- | --- |
| **Differentiated service delivery** | "I would be comfortable accessing PrEP even outside the clinic, in pharmacies and shops. That will make it even more accessible to people." (Migrant from Eastern Cape) |
| **Anonymity** | "Yes I am comfortable [testing for HIV] in Johannesburg and free. There are no chances of meeting somebody I know." (Migrant from Limpopo) |
| **PrEP information** | "PrEP need to be advertised more. It needs to get out there." (Migrant from KwaZulu-Natal) |
| **Social support** | "At the shelter there is tons of us. There are ladies and guys. There is six of us in a room. They are my new family." (Migrant from Limpopo) |
| **Masculinity** | "I sit down and talk to myself and tell myself that I am a man and that I can face any challenge." (Migrant from Limpopo) |
| **Positive coping skills** | "In the past I was very scared to test. But I have matured and I always want to go and test for HIV. When I go I come back happy. I always expect anything and I have courage to deal with whatever results I get. I think I am comforted by the fact that HIV can be managed by treatment." (Migrant from KwaZulu-Natal) |

Social support from other men surfaced as an important resource for some participants, including through religious organizations, bars, and sporting activities. At the homeless shelter, participants described how ten men in a room together would “share stories every night before we go to bed,” while others had support from family members or friends already in Johannesburg. In contrast to participants who described their sense of masculinity as a deterrent to care seeking (e.g., considering their wife’s HIV status as a proxy for their own), for other men, their sense of their role as a man motivated HIV testing and prevention. For example, men described a sense of responsibility for their family: “You don’t want to die and leave your family behind.” Others drew on their sense of masculinity or felt compelled to test for the sake of their partner: “I am the man and I have to protect my partner.” Lastly, some participants described positive coping skills which help them to feel “courage to deal with whatever results” from HIV testing.

## Discussion

Our findings of barriers to and opportunities for HIV testing and PrEP for internal migrant men in Johannesburg revealed themes common to men generally as well as themes exacerbated or unique to men who migrate internally within South Africa across all levels of influence (individual, social, structural), as shown in Figure 1. Similar to prior research, we found that substantial fears of an HIV-positive diagnosis^47–49^ and anticipated stigma^53,54^ remain major barriers to seeking HIV testing for men. Building on this research, our data highlight that men worry that a positive HIV diagnosis on top of pre-existing livelihood stressors would overwhelm their ability to cope and thus their mental wellbeing. Unemployment rates in South Africa are greater than 30%,^55^ and studies have shown the stressful toll that unemployment takes on South African men, leading not only to financial strain but also lower self-esteem and decreased social status.^56^ This suggests the need for interventions for men which address both livelihood stressors and coping skills. For example, a livelihood support intervention among persons living with HIV in Kenya showed evidence of mental health improvements,^57^ and the MenStar Strategy to engage men in HIV care in South Africa includes a focus on building coping potential.^58^ However, problematic alcohol use among men – prevalent in resource-constrained settings in South Africa and often the foundation of social gatherings^59^ – further complicates this picture as alcohol use may be associated with HIV risk behaviors.^60^ Efforts to address alcohol use and HIV risk have had mixed success, with evidence showing that social barriers (e.g., social norms around substance use) and structural level barriers (e.g., unemployment, poverty, violence, limited clinic resources) constrain long-lasting behavior change.^61,62^

**Figure 1.**
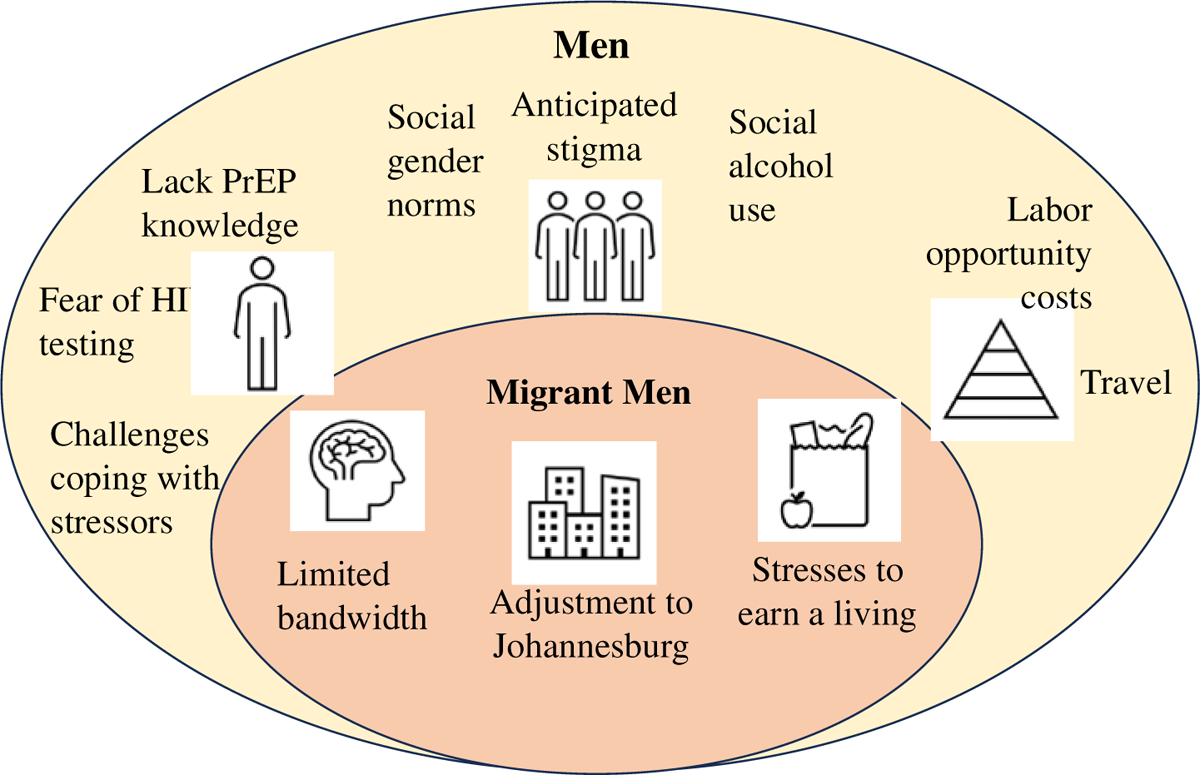
Barriers to HIV testing and prevention for men and internal migrant men in Johannesburg. In the yellow oval in the figure below, we show themes found to be common to men more generally, including migrants, organized by individual, social and structural levels of influence. In addition, we found themes which were heightened for migrant men, shown in the pink oval.

Building on these barriers for men, our data uncovered challenges that are exacerbated for and/or unique to South African migrants. Migrants may have a higher likelihood of life stressors, given the conditions that prompted their move as well as their near-universal need to find employment. Migrants are also faced with numerous additional social and structural stressors by virtue of needing to start afresh in a new city, including securing housing, navigating a new health system, building a new social network, prioritizing physical safety, and adjusting to the fast pace of life in Johannesburg. Each stressor takes up some degree of an individual’s “cognitive bandwidth,” which encompasses both cognitive capacity (psychological mechanisms which allow for reasoning, problem solving, and retaining information) and executive control (ability to plan, manage attention, and control actions).^63^ Each individual has a finite amount of cognitive bandwidth. An accumulation of stressors can tax this bandwidth and impede cognitive functioning, making it more difficult to manage immediate stressors, let alone to process new information (e.g., about PrEP) or prioritize HIV testing or preventative care.^63–65^ Similarly, research among international migrants has found that high levels of unemployment and competing life priorities, especially in the early phase of resettlement, can make it harder for migrants to seek preventative care.^66^ In addition to interventions focusing on livelihood and coping strategies, interventions which help migrants to adjust to life in their new city and navigate available health resources may also facilitate care engagement. For example, community navigator health workers in North America have been used to offer culturally tailored educational and communication support to guide migrants in overcoming barriers to healthcare.^67,68^

Despite these challenges, our data identified opportunities to engage migrants in HIV care, particularly for new arrivals to Johannesburg. As is the case in many countries worldwide, rural communities in South Africa often have less access to healthcare as compared to access in urban centers,^69^ and fears of unintentional disclosure may be heightened in rural clinics where seeking care means being seen by known community members.^70^ As our data showed, Johannesburg offers migrants greater opportunities to not only seek convenient, anonymous care but also to learn about HIV prevention options, such as PrEP. Similarly, research among international migrants has shown that many first learn about PrEP in their destination countries.^66^ In addition, migration may lessen the influence of family and culture norms from one’s home of origin, e.g. social constructs of gender which may limit men’s care-seeking.^53^ While some of our participants reported either social isolation or relying on pre-existing family members in Johannesburg, others described information sharing and new bonds that formed between themselves and other migrant men who were new to the city, especially for participants in the shelter. Thus, particularly for newcomers to Johannesburg, there may be an opportunity to provide easily accessible HIV services to migrant men at a time when they may be less constrained by prior social obligations and freer to adopt new social norms and behaviors.^71^

This study must be interpreted within limitations of this work. Many participants in our sample were unaware of PrEP, and none reported using it, similar to other research on barriers and facilitators of PrEP uptake for men in sub-Saharan Africa.^20,72,73^ Thus, it is possible that PrEP may have theoretical appeal in the context of first hearing about it, and social desirability bias may impact participants’ enthusiasm for PrEP as part of a research study. Barriers to PrEP use may emerge more prominently for men with experience using it. For example, accessibility issues were found to impede continuation of PrEP among women who previously used it as part of a study in South Africa.^74^ In addition, given the nature of this qualitative study and our focus on two recruitment sites in Johannesburg, the results cannot be generalized to all migrant populations within or outside of Johannesburg, nor can we statistically analyze differences between different sub-populations, e.g., newcomers versus longer-term migrants. Rather, we aimed to shed light on mechanisms underlying engagement in HIV testing and prevention to deepen our understanding of experiences for a random sample of internal migrants seeking shelter and work in the city. Many of the themes that we uncovered were similar to those found previously for men more generally, suggesting that strategies to engage migrant men (e.g., workplace HIV testing and PrEP availability) may have benefits for other men as well. Future research focusing on international migrants in South Africa is also warranted to understand the factors influencing their HIV care engagement.

## Conclusions

Migrant men come to Johannesburg to find work, but their struggle to survive without reliable income causes daily stress. Despite caring about their health, constraints on their time and cognitive bandwidth limit their ability to seek health services, particularly given fears of testing positive for HIV, anticipated stigma, and limited knowledge about the opportunity for PrEP should they test negative. Yet Johannesburg also presents opportunities for HIV testing and PrEP use for migrant men who perceive greater availability and anonymity of HIV information and services in the city as compared to rural homes of origin, and also more willingness to adopt new beliefs and behaviors. Bringing HIV services to migrant men at community sites may ease the burden of accessing these services. Including PrEP counseling and services alongside HIV testing may further encourage men to test, particularly if integrated into counseling on livelihood and coping strategies, as well as support for navigating health services in Johannesburg.

## Data Availability

All data produced in the present study are available upon reasonable request to the authors.

